# Cardiovascular Eligibility Criteria and Adverse Event Reporting in Cancer Therapy Trials of Combined Immune Checkpoint and VEGF Inhibitors: A Systematic Review

**DOI:** 10.1101/2023.07.14.23292585

**Authors:** Stephen Rankin, Benjamin Elyan, Robert Jones, Balaji Venugopal, Patrick B Mark, Jennifer S Lees, Mark C Petrie, Ninian N Lang

## Abstract

**Background:** Combination therapy with immune checkpoint inhibitors (ICI) and vascular endothelial growth factor inhibitors (VEGFI) has improved cancer outcomes and are increasingly common treatment regimens. These drug classes are associated with cardiovascular toxicities when used alone but heterogeneity in trial design and reporting may limit knowledge of toxicities in people receiving these in combination. Our aims were to assess consistency and clarity in definitions and reporting of cardiovascular eligibility criteria, baseline characteristics and cardiovascular adverse events in ICI/VEGFI combination trials.

**Methods:** Systematic review of phase II-IV randomised controlled trials of ICI/VEGFI combination therapy for solid organ cancer. We assessed trial cardiovascular eligibility criteria and baseline cardiovascular characteristic reporting in trial publications. We also examined cardiovascular adverse events definitions and reporting criteria.

**Results:** Seventeen trials (10,313 participants; published 2018-2022) were included. There were multiple cardiovascular exclusion criteria in 15 trials. No primary trial publication reported baseline cardiovascular characteristics. Thirteen trials excluded people with prior heart failure, myocardial infarction, hypertension or stroke. There was heterogeneity in defining cardiovascular conditions. Grade 1-4 cardiovascular adverse events were reported when incidence was ≥5-25% in 15 trials. Nine trials applied a more sensitive threshold for reporting higher grade AEs (severity grade ≥3 or serious AE). Safety follow up was shorter than efficacy follow up. Incident hypertension was recorded in all trials but other cardiovascular events were not consistently reported. Myocardial infarction was only reported in four trials and heart failure was reported in three trials. No trial specifically noted the absence of events. Therefore, in trials that did not report CVAEs, it was unclear whether this was because CVAEs did not occur. AE reporting and classification were by the investigator without further adjudication in 16 trials and one trial had an independent CVAE adjudication committee.

**Conclusions:** In ICI/VEGFI combination trials, there is heterogeneity in cardiovascular exclusion criteria, reporting of baseline characteristics and lack of reporting of cardiovascular adverse events. This limits optimal understanding of the incidence and severity of events relating to these combinations. Better standardisation of these elements should be pursued.

**Clinical Perspective:** *What is new?:*

- Immune checkpoint inhibitors (ICI) and VEGF inhibitors (VEGFI) are vital anti-cancer drugs but are associated with cardiovascular (CV) adverse events when given in isolation.
- VEGFI and ICI are now frequently used in combination, often in patients with pre-existing cardiovascular disease, but trial data to guide their use in such patients is limited.
- This systematic review of pivotal ICI/VEGFI trials identified heterogeneity in trial exclusion for pre-existing cardiovascular disease, reporting of CV baseline characteristics as well as in definitions and reporting of CV adverse events.

*What are the clinical implications?:*

- ICI/VEGFI oncology trial design and reporting methodology limits optimum understanding of adverse cardiovascular effects associated with ICI/VEGFI combination therapy, and these concerns may be more, or less, common than currently feared.
- Standardised cardiovascular eligibility criteria and adverse event recording would allow meta-analysis for more accurate assessments of adverse cardiovascular effects of ICI/VEGFI combination therapy.
- These observations and conclusions are relevant to the design and reporting of the majority of oncology drug trials and have implications to almost all anti-cancer therapeutic classes.

## INTRODUCTION

There is a high prevalence of cardiovascular disease (CVD) in people with cancer[1]. The incidence of cardiovascular (CV) events, such as myocardial infarction (MI) and ischaemic stroke, is higher in people with cancer than it is in people without cancer [2,3]. As clinical outcomes for people diagnosed with cancer have improved considerably over the past two decades, the competing risks from cardiovascular comorbidity and mortality have gained increasing relevance[4,5]

Therapies such as immune checkpoint inhibitors (ICI) and vascular endothelial growth factor inhibitors (VEGFI) have improved cancer outcomes for people with a variety of tumour types[6–8]. When used alone, ICIs are associated with a range of CV adverse effects (CVAE) including myocarditis, MI and ischaemic stroke[4,5,9–13]. VEGFI are also associated with a range of cardiovascular toxicities particularly hypertension, as well as left ventricular systolic dysfunction (LVSD), heart failure (HF) and atherothrombotic sequelae including MI and stroke[14–19].

The use of ICI and VEGFI in combination is now a common treatment regimen licensed in various cancer types, including melanoma, renal, cervical, and endometrial cancer[20]. This is a consequence of successful trials of combinations of ICI/VEGFI conducted over the last five years with more than ninety clinical trials of ICI/VEGFI combination regimes ongoing[8,21]. Six combination ICI/VEGFI treatments are currently approved by the Food and Drug Administration (FDA)[20]. Given the CV adverse effects seen with each of these drugs in isolation, understanding the potential for an increased incidence of these effects when the drugs are combined is of major importance.

There is limited understanding of the extent to which pre-existing CVD increases the risk of ICI/VEGFI cardiovascular toxicity. To understand these issues, it is imperative to know the proportion of people with pre-existing CVD who are excluded from trials. Understanding and limiting heterogeneity between trial populations is required for subsequent robust meta-analysis of CVAEs. Furthermore, consistency and clarity of definitions and trial publication reporting of CVAEs is fundamental to achieving these aims.

We conducted a systematic review of randomised controlled trials of ICI/VEGFI combination therapy in patients with cancer. Our primary interests were trial cardiovascular exclusion criteria and the heterogeneity of these between trials. We also examined methods by which adverse events (AEs) were defined, adjudicated and reported in trial results publications.

## METHODS

This systematic review protocol was registered on PROSPERO (CRD42022337942) and used the Preferred Reporting Items for Systematic reviews and Meta-Analyses (PRISMA) statement guidance[22]. We used the Population, Intervention, Comparison, Outcome (PICO) criteria for inclusion (Supplementary Table 1).

### Study eligibility criteria

A systematic search of the literature was conducted to identify clinical trials of combination ICI/VEGFI therapy. We included any trial conducted in an adult population with any solid organ cancer who received combination ICI/VEGFI therapy in either the intervention or the control arm. ICIs and VEGFI that were not approved by the FDA for use as an anti-cancer treatment at the time of data extraction were excluded. Trials using only single dosing or sequential (non-concurrent) ICI/VEGFI therapy were excluded.

### Inclusion and exclusion criteria

We included all phase II-IV randomised controlled trial with a minimum population of 20 participants that was published at time of extraction. Non-randomised controlled trials, meta-analyses, review articles, commentaries, subsequent therapy analyses, cost effectiveness analyses, published abstracts, patient reported outcomes, subgroup analyses and retrospective analyses were excluded. If two published articles reported data from the same patient group, such as subgroup analyses and extended follow up analyses, the original article was used.

### Outcomes

Key trial characteristics, trial eligibility criteria and exclusion criteria relating to cardiovascular disease were collected. Trial design characteristics relating to the assessment, adjudication of CVAEs and the extent of reporting of these within the published article were recorded. Data were extracted from the original publication, supplemental material and available protocols from the journal website. Trial registration numbers, identified from the publication, were used to search relevant clinical trial platforms to ensure all relevant publicly available protocol data were identified if they were not available from the publication.

### Cardiovascular adverse events

An AE was defined as a CVAE if it was recorded as a cardiac disorder under the Common Terminology Criteria for Adverse Event (CTCAE) criteria[23]. CTCAE criteria grades AE severity on a scale of 1 to 5. Grade 1 events are considered ‘mild’ and Grade 2 ‘moderate’. Grade 3 events are considered to be ‘severe or medically significant but not immediately life-threatening’ while Grade 4 events reflect those with life-threatening consequences. Death is recorded as Grade 5. If the AE was not recorded as a cardiac disorder by CTCAE but fulfilled any pre-specified trial criteria for cardiovascular and stroke endpoints for clinical trials, based on FDA endorsed Hicks’ criteria (such as ‘sudden death’), it was also classified as a CVAE[24].

### Search strategy

The search was conducted on MEDLINE, Embase and Cochrane Library in May 2022. The search terms are included in the Appendix. Duplicates were removed. Relevant articles were identified by two independent reviewers (B.E and S.R). Disagreements were resolved by consensus with a third reviewer (J.S.L).

## RESULTS

The search identified 4893 references which were screened (Figure 1). The final analysis included 17 randomised controlled trials with a total of 10313 participants, published between 2018 and 2022 (Table 1). Twelve were Phase III trials (9687 participants, 94%) and five were Phase II (626 participants, 6%). There were eight different combinations of ICI and VEGFI used. Atezolizumab with bevacizumab was the most common combination (Supplementary Table 3) used in six trials, (4357 participants, 42%).

**Figure 1:**
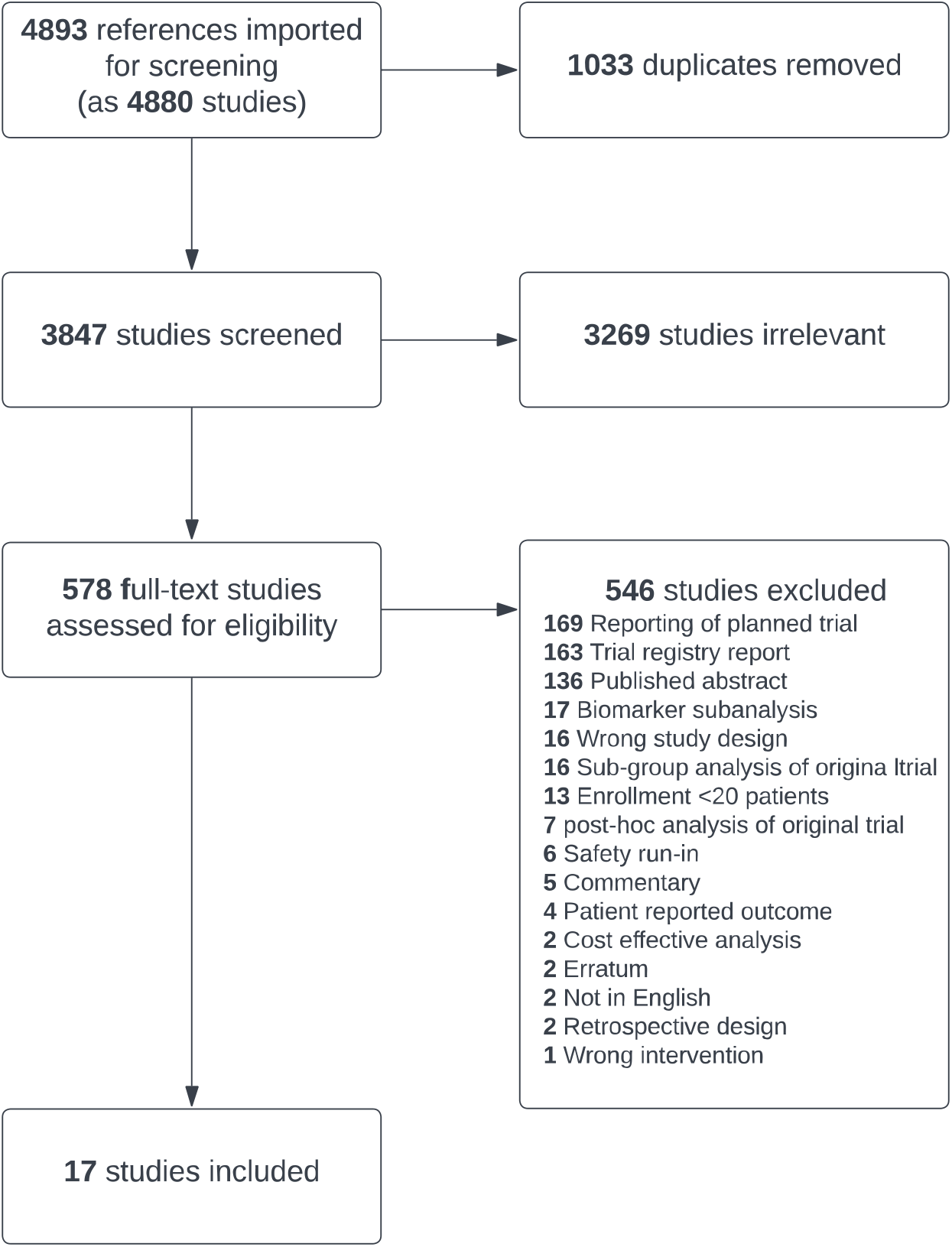
PRISMA Diagram.

**Table 1.**
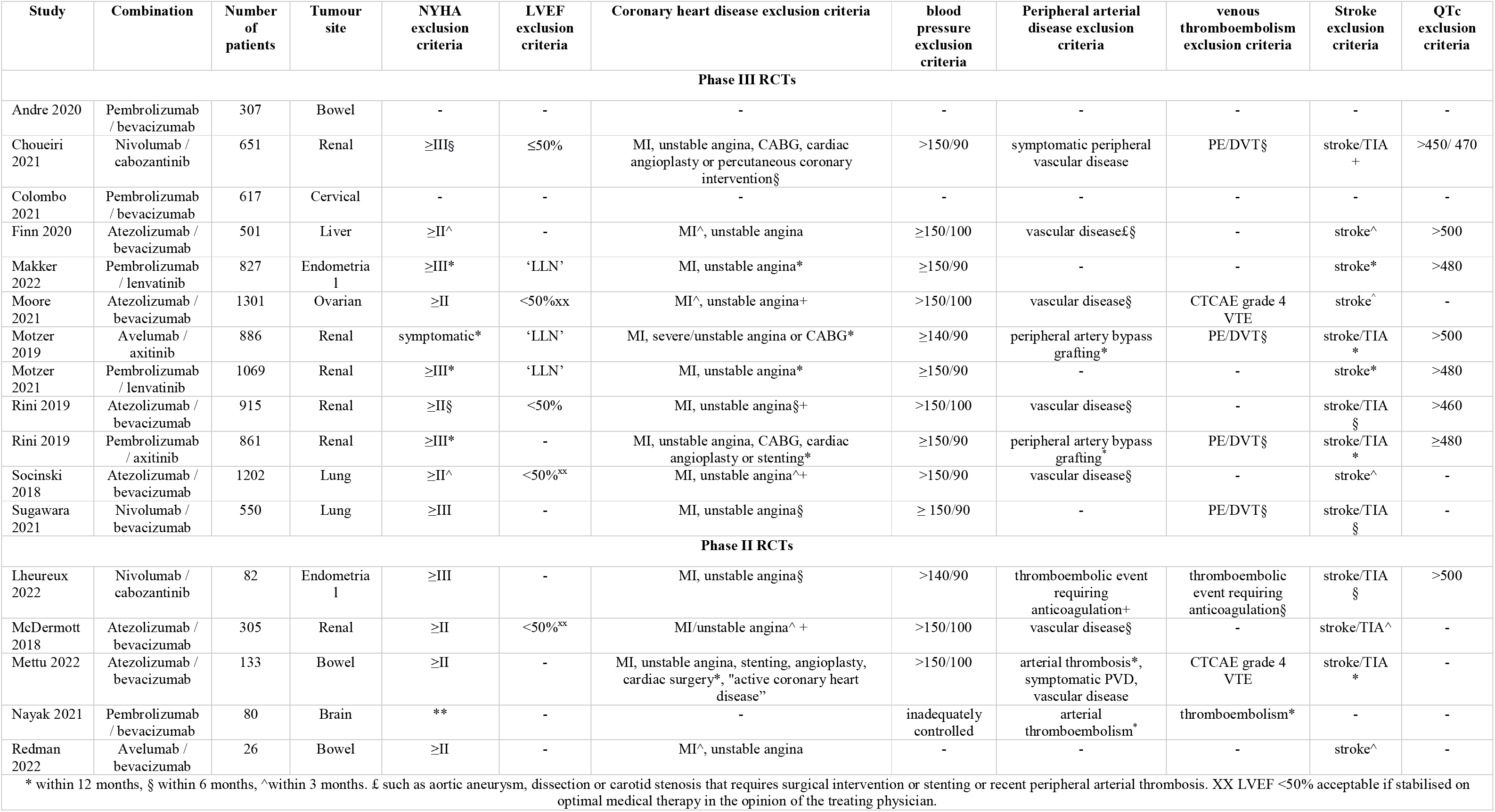
Randomised Controlled Trials of ICI/VEGFI combination therapy - Exclusion criteria.

**Table 2.**
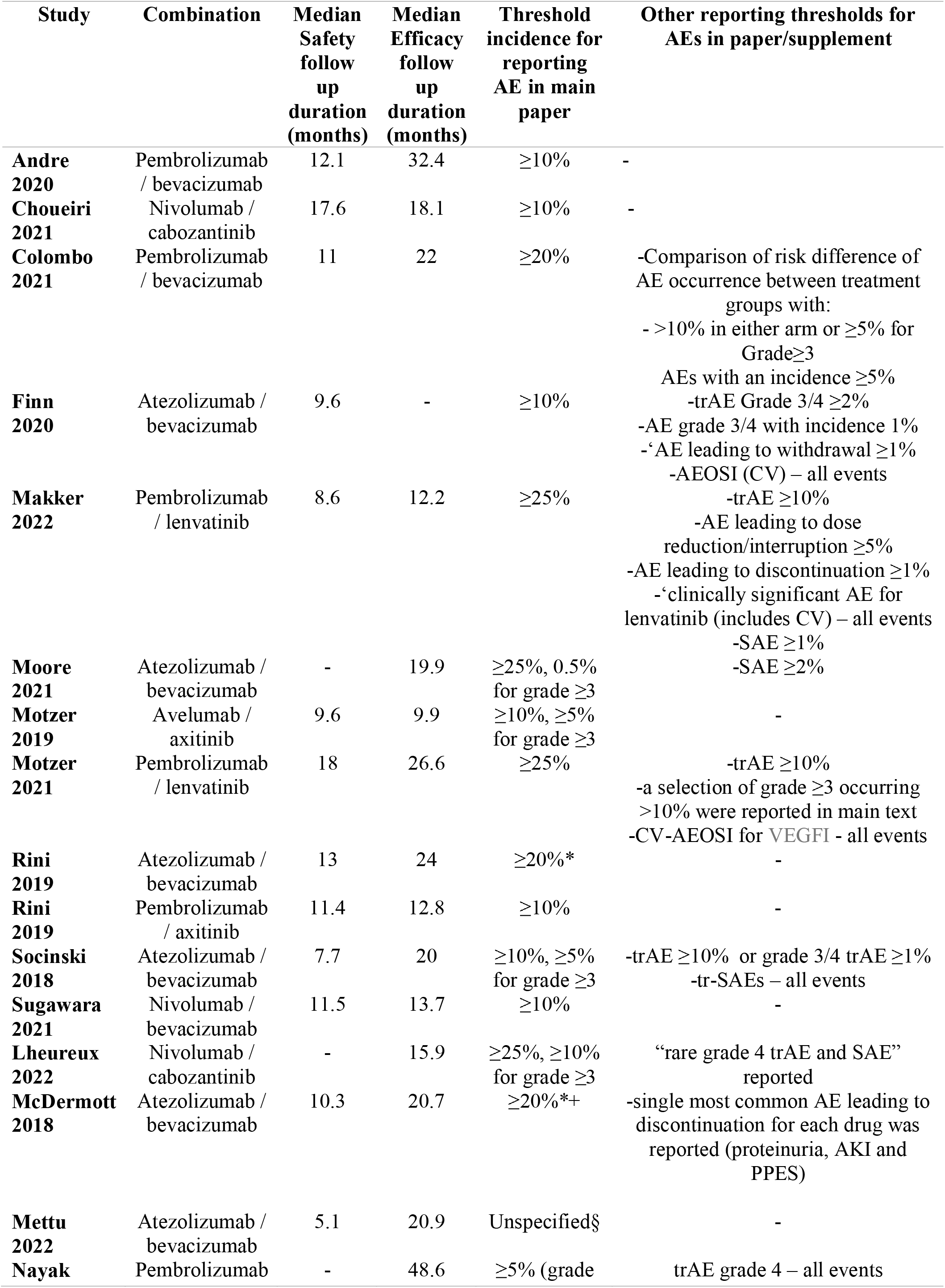
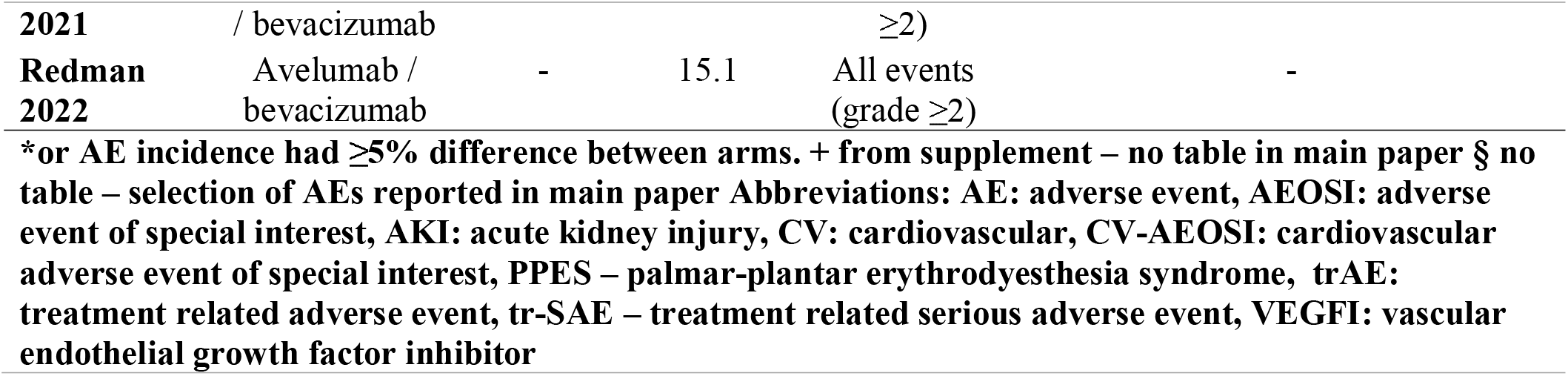
Reporting of CTCAE Grade 1-4 Adverse Events.

### Cardiovascular Eligibility Criteria

Eligibility criteria were available for all 17 trials. CVD trial exclusion criteria were broad with heterogenous definitions (Figure 2). Fifteen trials (9389 participants, 91%) had multiple CV exclusion criteria. There were specific exclusion criteria for people with prior HF, MI/unstable angina, hypertension and stroke in 13 trials (9283 participants, 90%). A further two trials (106 participants) had a general exclusion criterion of ‘clinically significant cardiovascular disease or impairment’. Two trials (924 participants) did not explicitly exclude people on the basis of prior CVD but had a general criterion excluding those with ‘a relevant prior condition which may affect the results of the trial’. The interpretation of these more generic criteria was left at the discretion of the investigator.

**Figure 2:**
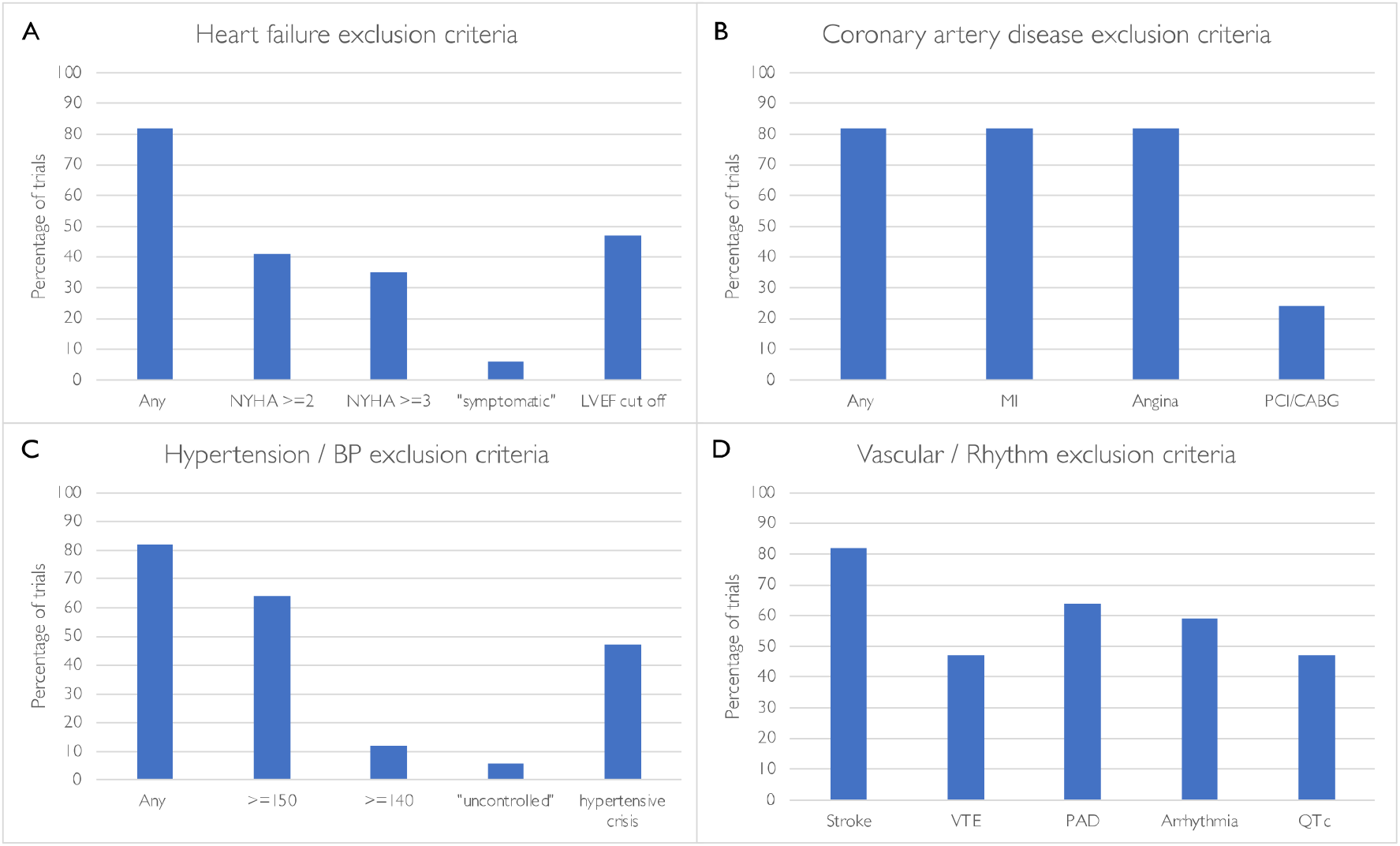
Cardiovascular exclusion criteria in ICI/VEGFi combination therapy trials. Abbreviations: NYHA: New York Heart Association, LVEF: left ventricular ejection fraction, MI: myocardial infarction, PCI: percutaneous coronary intervention, CABG: coronary artery bypass grafting, BP: blood pressure, VTE: venous thromboembolism

In the 12 trials reporting eligibility data prior to enrolment, 31% (3905 people) were ineligible. Only one paper reported reasons for screen failure and in that publication, 10% of those ineligible were excluded because of cardiovascular exclusions (PE/DVT, hypertension, QTc and ‘cardiovascular conditions’).

#### Heart Failure & Left Ventricular Systolic Dysfunction (LVSD)

Of the 14 trials (9309 participants, 88%) with specific exclusions for people with heart failure, seven excluded those with New York Heart Association (NYHA) ≥II, six excluded NYHA ≥III and one trial excluded ‘symptomatic’ patients (Table 1). Eight of the trials’ heart failure exclusions specified heart failure within a varying time frame prior to enrolment, ranging between 3-12 months prior to screening.

People with reduced left ventricular ejection fraction (LVEF) were excluded from eight trials (7156 participants, 69%): five excluded those with LVEF <50% (although three of these accepted LVEF <50% if the participant was ‘stable on a medical regimen that was optimised in the opinion of the physician’) and three excluded people with LVEF less than the ‘lower limit of normal’ of the ‘institutional normal range.’ Only four trials (3433 participants, 33%) mandated echocardiography before enrolment for all participants. Three other trials (1909 participants, 19%) mandated LVEF assessment prior to enrolment in specific circumstances (for patients with anthracycline exposure in one trial and, in if a patient had ‘cardiac risk factors or an abnormal electrocardiogram (ECG)’ in the remaining two).

There were exceptions to allow inclusion of participants with prior heart failure. In four trials, people with HF who did not meet pre-specified NYHA exclusion criteria, as well as people with LVEF <50%, were eligible to enrol provided they were on a stable regimen that was optimised in the opinion of the physician.

#### Coronary Artery Disease

The 14 trials with LVSD/heart failure exclusions also excluded patients with a history of recent MI/unstable angina (Table 1). The timeframe for exclusion of people with prior acute coronary syndrome varied from 3-12 months prior to screening. In addition to exclusions on the basis of acute coronary syndrome, four trials (2531 participants, 25%) also excluded people with coronary angioplasty, stenting or coronary artery bypass grafting (CABG) within 6-12 months prior to screening. In four trials, people known to have coronary artery disease (not otherwise meeting pre-specified coronary exclusions) were eligible for inclusion provided they were on a stable regimen that was optimised in the opinion of the physician.

#### Blood Pressure

Fifteen trials had a blood pressure (BP) or hypertension exclusion criterion (Table 1), most commonly excluding those with a systolic BP of 150mmHg and above (8315 participants, 81%). Two trials (106 participants) did not specify a BP cut-off but one trial excluded those with ‘inadequately controlled hypertension’ or a history of hypertensive encephalopathy/crisis. The second trial did not have a specific BP cut off but excluded participants randomised to receive bevacizumab if they had a previous history of hypertensive emergency or hypertensive encephalopathy. Any prior history of hypertensive encephalopathy or crisis was an exclusion in eight trials (4463 participants, 43%).

#### Stroke

Previous ‘cerebrovascular accident’ (CVA) or transient ischaemic attack within 3-12 months of screening was an exclusion criterion in 14 trials (9309 participants, 90%).

#### Arterial disease

Arterial vascular disease, such as aortic aneurysm requiring surgical repair, peripheral artery bypass grafting, peripheral arterial thrombosis in the 6-12 months prior to screening, was an exclusion criterion in 11 trials (6917 participants, 67%). There was heterogeneity in the definition of arterial disease, varying from those with surgical intervention (peripheral

artery bypass grafting) or those with any form of intervention or arterial thrombus in the preceding 6-12 months. Symptomatic PVD was an exclusion criterion in two trials.

#### Venous Thromboembolism

“Prior pulmonary embolism (PE) or deep vein thrombosis (DVT)” was an exclusion criterion in eight trials (4544 participants, 44%, Table 1), three of which had a time limit of exclusion to within the preceding 6 months. Venous thromboembolism (VTE) exclusion criteria were defined as either ‘PE/DVT within the preceding 6 months’ (one trial allowed those with recent PE/DVT provided they were stable on low-molecular weight heparin for six weeks) or a previous ‘CTCAE grade 4 VTE’ in two trials.

#### QTc & Arrhythmia

Patients with arrhythmia were excluded from ten trials (6482 participants, 63%). ‘Unstable’ or ‘haemodynamically significant’ arrhythmia was the most common exclusion terminology but ‘grade ≥2,’ ‘uncontrolled’ arrhythmias, and “clinically significant arrhythmias” were used to define this in three trials. Eight trials (5792 participants, 56%) had an upper QTc limit for enrolment, between 450-500milliseconds. Only one trial used a different threshold for men and women.

#### Myocarditis

No trial specifically excluded those with previous myocarditis, however every trial excluded patients with recent or current use of corticosteroids or immunosuppression, or previous hypersensitivity to ICI.

### Reporting of Baseline Cardiovascular Characteristics

With the exception of smoking status, which was reported in two lung cancer trials, no trial reported baseline CV characteristics, such as the prevalence of previous MI, heart failure, LVSD, diabetes, dyslipidaemia or hypertension.

### Reporting of Adverse Events

All 17 trials reported adverse events using CTCAE definitions and severity grading. CTCAE Version 4 was used in 15 trials. AEs were reported by the site investigator with no central or CV specialist event adjudication in 14 trials and was not specified in the remaining three trials. One trial had an independent cardiovascular events adjudication committee. AEs were either reported as treatment related (trAE) or ‘AEs of any attribution.’ TrAEs (adjudicated by the investigator) were reported in all trials. AEs of any attribution were less commonly reported (11 trials, 7458 participants, 72%).

#### Duration of Adverse Event Reporting

Follow-up for CV events was shorter than the trial duration in all trials (Table 3). Follow up for CV events in five trials was ‘the duration of treatment plus 30 days after last dose.’ In nine trials follow up for CVAEs was ‘duration of treatment plus 30 days or the initiation of new anti-cancer therapy, whichever came first’. In three other trials follow up was of 90-100 days for both AE and serious AEs (SAEs). Ten trials had extended follow up for SAEs and AE of special interest (AEOSI), including CVAEs, ranging from 90-120 days. The follow up period was not specified in two trials.

#### Incidence Thresholds for Adverse Event Reporting

No phase III trial reported all CV events. Fifteen trials reported events when they reached a pre-specified incidence (Table 3). The most common threshold in the main paper was ≥10% in six trials (3756 participants, 36%) but higher reporting thresholds (incidence of ≥20-25%) were used in 5 trials (4307 participants, 42%). One phase II trial (26 participants, 0.25%) reported all CTCAE grade ≥2 treatment related AEs. A lower threshold specifically for reporting more severe AEs (CTCAE grade ≥3 or SAE) was used in nine trials (6565 participants, 64%) and that threshold ranged from ‘all events’ to 10%. Three trials (1633 participants, 16%) reported AEs under a variety of other specific circumstances with lower thresholds, such as ‘AEs leading to discontinuation’ (Table 3). Grade 5 AEs (deaths) were reported in all trials. Twelve trials (7854 participants, 76%) reported AE deaths regardless of relationship to treatment, where 5 trials (2459 participants, 24%) only reported treatment related AE deaths, adjudicated by the investigator.

### Cardiovascular events

No trial used the FDA-endorsed, standardised Hicks’ criteria for reporting of cardiovascular events[24]. With the exception of hypertension, which was reported in all trials, no trial explicitly stated the absence or occurrence of CVAEs. In trial manuscripts that did not report CVAEs other than hypertension, it was not clear whether this was because of a true absence of CVAEs or because of their occurrence with an incidence beneath a reporting threshold.

#### CV death

‘AE deaths of any attribution’ were reported in 11 trials (7203 participants) and 7 of these (4734 participants) reported the mode of AE death. In six trials (3110 participants), only deaths that were considered to be treatment related (adjudicated by the investigator) were reported.

Most frequently, CVAEs were described when associated with death. No trial reported total number of CV deaths. However, ten trials (7737 participants) reported AE death that would be categorised as CV death by Hicks’ Criteria (Figure 3).

**Figure 3.**
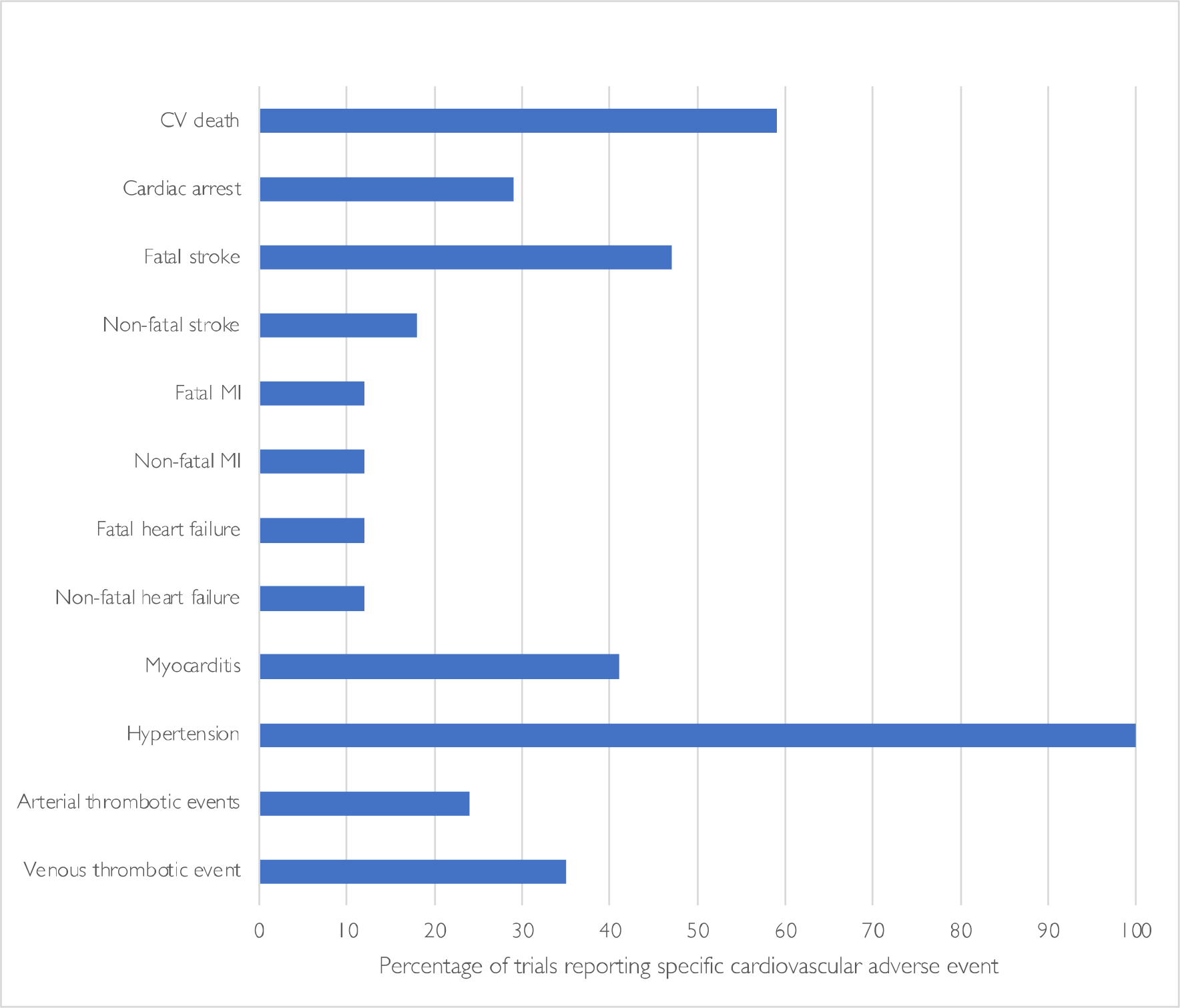
Proportion of trials reporting CV adverse events. Abbreviations: CV: cardiovascular, MI: myocardial infarction

#### Myocardial Infarction

MI was only reported in four trials (3181 participants, 31%), two of which only reported fatal MI (Figure 3). No trial reported if coronary revascularisation occurred.

#### Heart failure & LVSD

HF was reported in three trials (2564 participants, 25%), two of which reported one fatal case of HF. One trial reported one case of fatal cardiac failure and three cases of grade 1-2 ‘congestive cardiac failure’ defined by CTCAE Version 4. LVSD was also reported in three trials (3,098 participants, 30%). Two of the three trials that reported LVSD mandated echocardiography surveillance on treatment. Four trials (2913 participants, 28%) reported ‘peripheral oedema’.

#### Stroke

Stroke was reported in eight trials (5782 participants, 56%). Five trials (2778 participants, 27%) only reported the occurrence of fatal stroke. Ischaemic stroke was reported in five trials (4536 participants, 44%). Fatal ischaemic stroke occurred in four of these trials, three of which were only reported in supplementary data. Haemorrhagic strokes were reported in six trials (3864 participants, 38%) and five of these trials only reported fatal haemorrhagic strokes.

#### Myocarditis

Myocarditis occurred in seven trials (5309 participants, 52%). Fatal myocarditis was reported in two trials. No trial reported if myocarditis did not occur.

#### Hypertension

Hypertension was reported in all trials, defined by CTCAE. Posterior reversible encephalopathy syndrome (PRES) was reported in two trials (2271 participants, 22%). There were two reported deaths attributed to hypertension: one secondary to PRES and another death secondary to ‘uncontrolled hypertension’ adjudicated by the investigator.

#### Other thrombotic events

Venous thrombotic events were reported in six trials (5309 participants, 52%) but four of these only reported thrombotic events that resulted in death. Four trials (3599 participants, 35%) reported arterial thrombotic events and three trials (1944 participants, 19%) reported unspecified thromboembolic events.

#### Adverse Events of Special Interest (AEOSI)

AEOSI were collected in 15 trials (10205 participants, 99%), all of which included the collection of immune-related AE (irAE), including myocarditis. All 15 trials reported ir- AEOSI, with lower incidence thresholds (all ir-AEOSI events in 13 trials, >1% in the ICI arm in one trial, and unspecified in one trial).

CV-AEOSI were collected or reported in six trials (4717 participants, 46%). CVAEs were included in AEOSI lists in the protocol of five of these trials (3890 participants, 38%). The definition of these CV-AEOSI varied from ‘grade ≥2 cardiac disorders’ to more comprehensive lists detailing reporting of venous, arterial thromboembolism, LVSD, significant, arrhythmias and heart failure events. Only four trials reported CV-AEOSI, however three of these reported CV-AEOSI only in supplementary materials (Table 3). No trial specifically reported that an AEOSI did not occur.

## DISCUSSION

This systematic review of randomized trials of ICI/VEGFI combination therapy demonstrates heterogeneity in three key areas relevant to potential adverse cardiovascular effects of these important anti-cancer drugs. First, cardiovascular trial exclusion criteria are inconsistent between trials. Second, primary trial manuscript reporting of the prevalence of cardiovascular disease and risk factors in trial participants is variable and limited. Third, there is variation in methods, thresholds and follow-up periods for reporting and publication of adverse cardiovascular events associated with ICI/VEGFI combination therapy.

This review focuses specifically on trials of combined ICI and VEGFI therapies. Given that both ICI and VEGFI are associated with a range of adverse cardiovascular effects when given alone[10,25], the most robust assessment of these events is of even greater relevance and importance when the drugs are given in combination. Furthermore, it is imperative to understand the baseline cardiovascular characteristics of trial participants to gauge generalizability of trial outcomes, including adverse cardiovascular events, in the wider ‘non-trial’ population receiving these drugs in routine clinical practice[26]. Randomised trials of combined ICI/VEGFI were first reported in 2018 and therefore represent contemporary trial methodology[27,28]. A prior review of a broad range of anti-cancer agents, including conventional chemotherapeutics, examined cardiovascular adverse event reporting in cancer trials supporting FDA approval but this included trials conducted over 30 years ago and was prior to any FDA approval of combination therapy[29].

### Cardiovascular Trial Eligibility Criteria Heterogeneity

Our review identified that cardiovascular exclusion criteria were ubiquitous in these trials. We also identified substantial heterogeneity in the nature of these exclusion criteria and the use of potentially arbitrary CV definitions and exclusion thresholds. It is of note that the FDA recommend the avoidance of ‘unnecessarily restrictive eligibility criteria’ to maximise the generalizability of trial results to the patient population in whom the drug may be used in subsequent routine clinical practice[30]. This recommendation is made particularly to allow trials to inform the net risk/benefit profile. While we acknowledge that it may be appropriate to include some clinically-relevant cardiovascular eligibility criteria for trial safety reasons, standardisation of these criteria would allow easier translation of trial results to guide treatment decisions for patients treated in everyday practice. Indeed, CV disease and risk factor burden is high in patients with cancer [1,31,32]. Furthermore, while these trials were designed and powered to provide information on cancer treatment effects, potential safety signals may only become apparent when trial populations are combined for meta-analysis. Those insights are currently limited by heterogeneity in eligibility criteria.

### Baseline CVD and CVD Risk Factors in Trial Participants

Baseline cardiovascular characteristics, including cardiovascular risk factors or established CVD, were not reported in any primary trial publication. However, a secondary analysis of one trial did report the prevalence of baseline CV risk factors[33]. In that report, the prevalence of CV risk factors was low. Only 4% of people in the ICI/VEGFI arm had dyslipidaemia, 9.5% had diabetes and 3.2% had cerebrovascular disease[33]. In addition to potentially stringent trial eligibility criteria, trial recruitment bias toward inclusion of people with fewer comorbidities may contribute to a trial population who are not representative of the general population of patients with cancer in whom these drugs may ultimately be used. Observational data suggest that fewer SAEs occur in trial participants than expected when compared to matched patients in routine clinical care and multi-morbidity is associated with higher events[34]. Irrespective of these issues of eligibility and potential recruitment bias, the lack of data on baseline CV characteristics means that the baseline CV risk for patients in these trials is unknown. Inclusion of those with comorbidities, when assessed in non-cancer trials, only modestly affected completion of study enrolment, meaning there could be an increase in generalisability of trial data with minimal impact on trial completion[35]. Without this information, it is not possible to assess the degree to which pre-existing CVD or risk factors may potentiate adverse cardiovascular effects of ICI/VEGFI therapy. It also remains possible that an interaction between pre-existing CVD and adverse effects of ICI/VEGFI therapy is lower than might otherwise be expected. These insights are critical for providing patients with the best information relating to potential risks of treatment in the context of pre-existing CVD.

### Cardiovascular Adverse Event Description and Reporting

CVAEs were reported using CTCAE criteria in all trials and reporting of these was based upon incidence thresholds. The threshold that was required to be reached varied from 5-25% between trials. Furthermore, only four trials used a lower reporting incidence threshold for more severe (CTCAE grade ≥3). In addition to standardization of reporting methods, lowering the threshold or potentially removing this threshold for reporting in primary trial publications altogether would appear reasonable. While the signal to noise ratio of grade 1 and 2 events may mean that reporting on the basis of incidence thresholds could be appropriate, we would argue that reporting of all of the more severe adverse events may be justified.

Trial publication reporting of CVAEs, and the clarity of this, was variable. While many primary trial publications did not report the occurrence of CVAEs, they also did not explicitly state their absence. Reporting of AEs that were specifically considered to have been related to treatment (trAEs) was more frequent than reporting of AEs of any attribution. The assessment of CVAE ‘treatment-relatedness’ was by the local investigator and therefore this introduces bias and impedes transparent understanding of AE profiles. One trial included a pre-specified subgroup analysis of cardiovascular events in ICI/VEGFI therapy. In that analysis, the number of CV events was small but CVAE incidence was higher than reported in the primary manuscript [33].

All trials had longer follow-up for anti-cancer efficacy assessment than they did for collection of CVAEs. Given that the accrual of CVAEs might be expected to occur over a similarly more prolonged period, increasing follow-up duration for CVAEs would provide important information.

### Limitations

This systematic review has several limitations. We did not collect data on pre-trial safety data which may have influenced eligibility criteria. We also did not collect data on subgroup analysis and extended follow up papers which may have provided additional information on CV comorbidities and adverse effects. However, given that original trial manuscripts frequently inform drug licensing approvals we believe that our focus on these publications is particularly relevant. It is also possible that some safety data are still to be placed in the public domain and therefore not captured.

### Conclusion

This systematic review of randomized trials of ICI/VEGFI combination therapies has identified heterogeneity in trial cardiovascular eligibility criteria, limited trial manuscript reporting of the baseline cardiovascular characteristics in participants and heterogeneity in methods used for reporting of adverse cardiovascular events. These factors may have substantial impact on the ability to make accurate assessments, including meta-analyses, of the potential for cardiovascular adverse effects of these important anti-cancer therapies. While it is possible that cardiovascular adverse effects are under-appreciated, it is also possible that they may be less frequent than feared. With the rapid rise of combination ICI/VEGFI treatment regimens there is an urgent need to standardise these components and, in particular, to inform their use in patients who frequently have pre-existing cardiovascular disease.

## Supporting information

supplemental review material

supplemental publication material

## Data Availability

original data can be made available on request.

## Acknowledgements

None

## Funding

SR receives support through an unrestricted grant from Roche Diagnostics.

## Disclosures

Each author has completed the ICJME conflicts of interest form. N.N.L. reports research grants from Roche Diagnostics, Astra Zeneca and Boehringer Ingelheim as well as consultancy/speaker’s fees from Roche Diagnostics, Myokardia, Pharmacosmos, Akero Therapeutics, CV6 Therapeutics, Jazz Pharma and Novartis all outside the submitted work. Outside the submitted work, J.S.L. has received personal lectureship honoraria from Astra Zeneca, Pfizer and Bristol Myers Squibb. P.B.M. reports grants and personal fees from Boehringer Ingelheim; honoraria from Astrazeneca, GSK, Pharmacosmos, and Astellas, outside the submitted work. B.E has no conflicts of interest to declare. S.R. receives support through an unrestricted grant from Roche Diagnostics. Outside of the submitted work, BV receives: Consultancy or advisory role: Bristol Myers Squibb, EUSA Pharma, Merck Sharp & Dohme; travel/accommodation/expenses: Bristol Myers Squibb, EUSA Pharma, Ipsen; Research funding (institution): Bristol Myers Squibb, Exelixis, Ipsen, Merck Sharp & Dohme, Pfizer; Honoraria (self): Bristol Myers Squibb, Ipsen, Pfizer; Speaker bureau/expert testimony: Bristol Myers Squibb, Eisai, EUSA Pharma, Merck Serono, Merck Sharp & Dohme, Pfizer. R.J.J. reports grants from Astellas, Clovis, Exelixis, Bayer and Roche; honoraria from Astellas, Janssen, Bayer, Pfizer, Merck Serono, MSD, Novartis, Roche, Ipsen, Bristol Myers Squib. M.C.P reports grants from Boehringer Ingelheim, Roche, SQ Innovations, Astra Zeneca, Novarttis, Novo Nordisk, Medtronic, Boston Scientific, Horizon and Phramacosmos, all outside the submitted work; honoraria from Boehringer Ingelheim, Novartis, Astra Zeneca, Novo Nordisk, Abbvie Bayer, Takeda, Corvia, Cardiorentis, Pharmacosmos, Siemens and Vifor.

## Supplemental Material

**Supplementary Table 1: PICO Framework**

**Supplementary Table 2: List of VEGF inhibitors and immune checkpoint inhibitors & systematic review search terms**

**Supplementary Table 3: Summary of trial characteristics and exclusions**

**Supplementary Figure 1: Percentage of participants in ICI/VEGFi combination trials subject to cardiovascular exclusion criteria**

**Supplementary Figure 2: Percentage of participants in ICI/VEGFi combination therapy trials for whom cardiovascular adverse events were reported**

**Supplementary file: PRISMA checklist Supplementary file: PRISMA abstract checklist**

## Non-standard Abbreviations & acronyms

AE: adverse events
BP: blood pressure
CV: cardiovascular
CVAE: cardiovascular adverse events
CVA: cerebrovascular accident
CTCAE: Common terminology criteria adverse events
AEOSI: adverse events of special interest
CV-AEOSI: cardiovascular adverse events of special interest
HF: heart failure
ICI: immune checkpoint inhibitor
ir-AE: immune related adverse events
LVSD: left ventricular systolic dysfunction
MI: myocardial infarction
PE: pulmonary embolism
SAE: serious adverse event
trAE: treatment related adverse event
VEGFI: Vascular endothelial growth factor inhibitor

## Notes

### Clinical Protocols

https://www.crd.york.ac.uk/prospero/display_record.php?RecordID=337942

### Author Declarations

source data was openly available before initiation of the study and was identified using web platforms outlined in methods, such as pubmed and clinicaltrials.gov

