## supplemental publication material for "Cardiovascular Eligibility Criteria and Adverse Event Reporting in Cancer Therapy Trials of Combined Immune Checkpoint and VEGF Inhibitors: A Systematic Review"

**Supplementary Table 1: PICO framework table.**

Outcomes of interest using PICO framework and data points of interest

| | |
|---|---|
| Population | Inclusion criteria:<br>- adult population with cancer,<br>- all solid organ tumour sites.<br><br>Exclusion criteria:<br>- animal studies. |
| Intervention | inclusion criteria:<br>- trials that included the use of a VEGF-inhibitors (VEGFI), and/or in combination with immune checkpoint inhibitors (ICIs) class of drugs for the treatment of cancer,<br>- VEGFI with or without ICI used as either control arm or intervention arm of study,<br>- phase II, III and IV trials,<br>- randomised studies,<br>- published in English language,<br>- completed enrolment,<br><br>The specific intervention exclusion criteria are listed below:<br>- single dosing,<br>- sequential therapy rather than concurrent therapy,<br>- population of treatment group of <20 patients,<br>-non-randomised trials<br>- meta-analysis,<br>- review articles or commentaries,<br>- subsequent therapy analysis, |

<table>
<tr><td></td><td>- cost-effective analysis,<br>- published abstracts,<br>- patient reported outcomes,<br>- subgroup analysis,<br>- duplications,<br>- retrospective analysis.<br><br>Trials that have incomplete protocol and text will be included in the results as part of our objective is to identify the representation of patients with kidney disease in trial data.</td></tr>
<tr><td>Comparison</td><td>Not applicable</td></tr>
<tr><td>Outcome</td><td>We will collect the following data points from the extracted articles regarding the exclusion and representation of patients with kidney disease:<br>1. Are patients with any form of kidney disease excluded in design/eligibility criteria?<br>– Was marker of kidney function used as exclusion criteria<br>– Definition(s) of kidney function used<br>– Cut off value(s)<br>– Were alternative measure of GFR allowed (MDRD, CKD-EPI, urine creatinine collection, measured)?<br>2. Were patients with proteinuria excluded from the trial?<br>– Method(s) used as proteinuria screen<br>o Urinalysis<br>o Urine PCR/ACR<br>o 24 hour urine collection<br>– Cut off value for exclusion<br>3. Were patients with kidney/solid organ transplants excluded?</td></tr>
</table>

| | |
|---|---|
| | 4. Were patients on regular immunosuppression excluded?<br><br>5. Did patients have nephrectomy?<br><br>6. Did the trial analyse effect of benign kidney disease on primary outcome?<br><br>7. Is the kidney function of trial population available in baseline characteristics or supplementary materials?<br><br><br>We plan to collect similar exclusions of patients with cardiovascular disease:<br><br>1. Were patients excluded with cardiovascular disease?<br><br>2. Were patients excluded with the following diagnosis:<br><br>- Hypertension<br><br>- Coronary artery disease (CAD)<br><br>- Heart failure<br><br>- Cardiomyopathy<br><br>- Arrhythmia<br><br>- Valvular disease<br><br>- Thromboembolic disease<br><br>- Cerebrovascular disease<br><br>- Abnormal ECG<br><br>- Myocarditis<br><br>- Pericarditis<br><br>- Vasculitis<br><br>3. Are the diagnosis of cardiovascular disease of trial population available in baseline characteristics or supplementary materials?<br><br>4. Are cardiovascular adverse events reported in the published clinical trial or supplement? |

| | |
|---|---|
| | We will also collect the following information about the trial characteristics to analysis if this had an influence on the trial design or patient enrolment:<br>1. Is the trial ID number identifiable?<br>2. Is the trial name identifiable?<br>3. What is the trial design?<br>4. Was this a randomised control design?<br>5. Was this intervention or control?<br>6. Was there an active comparator?<br>7. Trial year published<br>8. Trial population size<br>9. Funding source<br>10. Cancer diagnosis<br>11. Is trial protocol available?<br>12. How were adverse events defined and were they adjudicated?<br>13. Is there a clear safety follow up period specified in the protocol?<br>14. Is data collected on adverse events of special interest?<br>15. Are adverse events of special interest defined in the protocol? |

**Supplementary Table 2: List of VEGF inhibitors and immune checkpoint inhibitors & systematic review search terms**

<table>
<tr><th colspan="2">VEGF inhibitors</th></tr>
<tr><th colspan="2">Tyrosine kinase Inhibitor</th></tr>
<tr><td>Apatinib<br>Axitinib<br>Brivanib alaninate<br>Cabozantinib<br>Cediranib<br>Dovitinib<br>Lenvatinib<br>Nintedanib</td><td>Pazopanib<br>Regorafenib<br>Sorafenib<br>Sunitinib<br>Tivozinib<br>Vandetanib<br>Vatalanib</td></tr>
<tr><th colspan="2">Monoclonal antibodies</th></tr>
<tr><td>Bevacizumab</td><td>Ramucirumab</td></tr>
<tr><th colspan="2">VEGF-Trap mediators</th></tr>
<tr><td colspan="2">Aflibercept</td></tr>
<tr><th colspan="2">Immune checkpoint inhibitors</th></tr>
<tr><th colspan="2">PD-1</th></tr>
<tr><td>Pembrolizumab<br>Nivolumab</td><td>Cemiplimab</td></tr>
<tr><th colspan="2">PD-L1</th></tr>
<tr><td>Atezolizumab<br>Avelumab</td><td>Durvalumab</td></tr>
<tr><th colspan="2">CTLA-4</th></tr>
<tr><td>Ipilimumab</td><td>Tremelimumab</td></tr>
<tr><th colspan="2">Search Terms</th></tr>
<tr><td colspan="2">((“Phase II” OR “Phase 2” OR "Phase III" OR "Phase 3" OR "Phase IV" OR "Phase 4") AND (Ipilimumab OR Tremelimumab OR Atezolizumab OR Avelumab OR Durvalumab OR Pembrolizumab OR Nivolumab OR Cemiplimab) AND<br>(Apatinib OR Axitinib OR Bevacizumab OR Aflibercept OR 'Brivanib alaninate' OR Cabozantinib OR Cediranib OR Dovitinib OR Ramucirumab OR Lenvatinib OR Nintedanib OR Pazopanib OR Regorafenib OR Sorafenib OR Sunitinib OR Tivozanib OR Vandetanib OR Vatalanib))</td></tr>
</table>

Abbreviations: VEGF, Vascular endothelial growth factor; PD-1, Programmed cell death protein-1; PD-L1, Programmed death ligand-1; CTLA-4, Cytotoxic T-lymphocyte protein-4

### Supplementary Table 3. Summary of trial characteristics and exclusions

| | Number of trials n (%) | Number of patients n (%) |
|---|---|---|
| Total (randomized controlled trials) | 17 | 10313 |
| **Tumour type** | | |
| Renal | 6 (35) | 4687 (45) |
| Gynaecological (endometrial, ovarian, cervical) | 4 (24) | 2827 (27) |
| Lung | 2 (12) | 1752 (17) |
| Liver | 1 (6) | 501 (5) |
| Bowel | 3 (18) | 466 (5) |
| Brain | 1 (6) | 80 (1) |
| **Trial Phase** | | |
| III | 12 (71) | 9687 (94) |
| II | 5 (29) | 626 (6) |
| **ICI and VEGFi combination** | | |
| Atezolizumab & bevacizumab | 6 (35) | 4357 (42) |
| Pembrolizumab & Lenvatinib | 2 (12) | 1896 (18) |
| Pembrolizumab & bevacizumab | 3 (18) | 1004 (10) |
| Avelumab & axitinib | 1 (6) | 886 (9) |
| Pembrolizumab & axitinib | 1 (6) | 861 (8) |
| Nivolumab & cabozantinib | 2 (12) | 733 (7) |
| Nivolumab & bevacizumab | 1 (6) | 550 (5) |
| Avelumab & bevacizumab | 1 (6) | 26 (0.3) |

**Supplementary Figure 1: Percentage of participants in ICI/VEGFi combination trials subject to cardiovascular exclusion criteria**

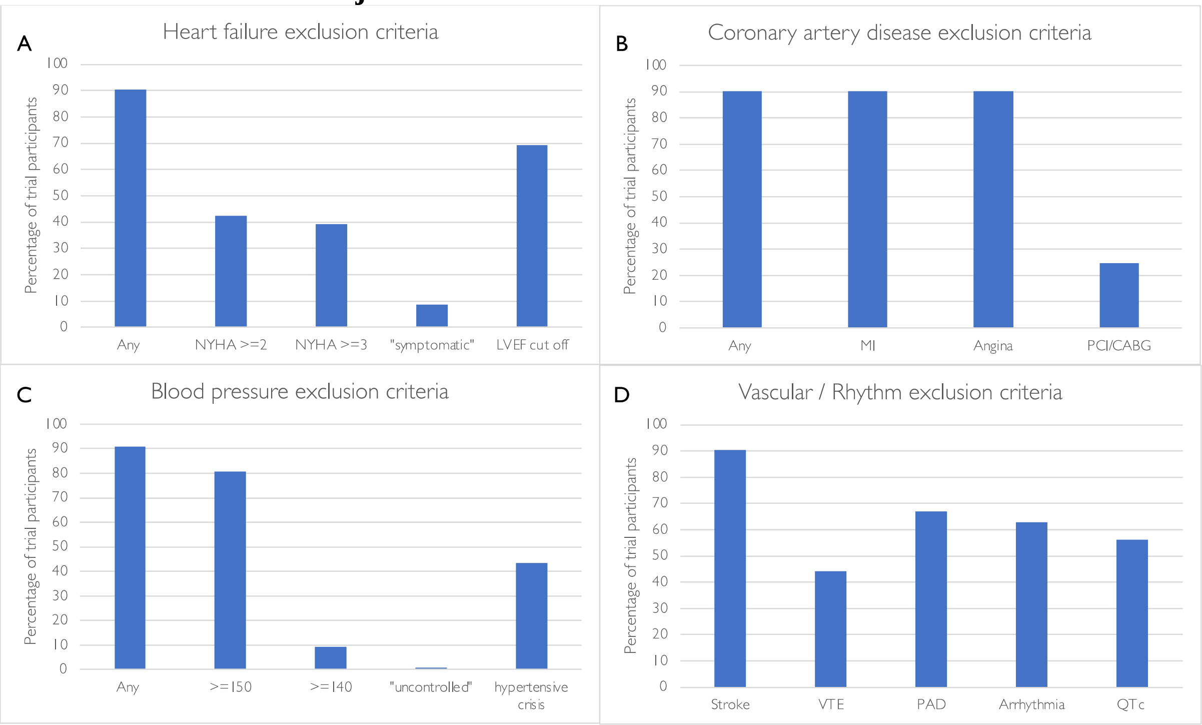

**Supplementary Figure 2: Percentage of participants in ICI/VEGFi combination therapy trials for whom cardiovascular adverse events were reported**

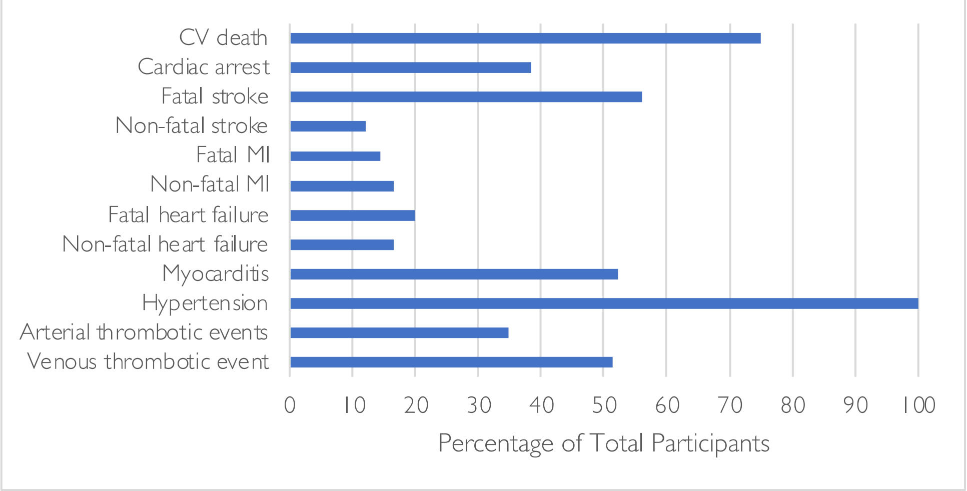
